# Hepatitis B infection is causally associated with extrahepatic cancers: a Mendelian randomization study

**DOI:** 10.1101/2021.03.13.21253528

**Authors:** Abram Bunya Kamiza, Segun Fatumo, Mwiza Gideon Singini, Chih-Ching Yeh, Tinashe Chikowore

## Abstract

Overwhelming evidence suggests that chronic hepatitis infection is associated with extrahepatic cancers. However, uncertainties exist about this association as much of the current evidence evolve from observational studies which are susceptible to confounding. We performed two-sample Mendelian randomization (MR) to explore the causal associations between chronic hepatitis infection and extrahepatic cancers. Genetic variants associated with chronic hepatitis B virus (HBV) infection and chronic hepatitis C virus (HCV) infection were identified from a large genome-wide association study. Summary level data for cancer of the biliary tract, cervix, colorectum, endometrium, esophagus, gastric, liver, lung, ovary and pancreas were obtained from the Biobank Japan. Using the inverse variance weighted method, we found chronic HBV infection to be causally associated with gastric cancer (odds ratio [OR] = 1.19 and 95% confidence interval [CI] = 1.13-1.25, P-value = 0.001) and lung cancer (OR = 1.21, 95% CI = 1.14-1.28, P-value = 0.001). Moreover, chronic HBV infection (OR = 1.34, 95% CI = 1.17-1.53, P-value = 0.007) and chronic HCV infection (OR = 2.75, 95% CI = 2.21-3.42, P-value = 0.0008) were all causally associated with liver cancer, supporting a well-established association between chronic hepatitis infection and liver cancer. In conclusions, our MR findings revealed that chronic HBV infection is causally associated with extrahepatic cancers including gastric and lung cancers.

## Introduction

Globally about 360 million people are chronically infected with hepatitis B virus (HBV) and hepatitis C virus (HCV) [1, 2]. Overwhelming evidence indicates that chronic hepatitis infection is the risk factor for liver cancer [3–5]. Further evidence suggests that chronic hepatitis infection is associated with extrahepatic cancers [6–10]. Moreover, our previous analysis revealed that chronic HBV infection is associated with cancer of the liver, pancreas, kidney, colorectum, ovary, non-Hodgkin’s lymphoma, gallbladder and extrahepatic bile duct whilst chronic HCV infection was associated with cancer of the liver, ovary, non-Hodgkin’s lymphoma, gallbladder and extrahepatic bile duct [11]. However, uncertainties exist about the association between chronic hepatitis infection and extrahepatic cancers as much of the current evidence evolve from observational studies [6–11], which are susceptible to confounding. Hence this association needs further evaluation using a different approach.

Mendelian randomisation (MR) is an analytical approach that uses genetic variants as instrumental variables to infer the causality of exposure to an outcome [12]. The MR is based on the idea that the inheritance of alleles are independent [13], thus suggesting that the exposure predicted by the genetic variants are independent of other exposures, thereby reducing biases commonly observed in observational studies. Unlike observational studies, MR analyses are not confounded by some unmeasured factors owing to the random independent segregation of alleles during meiosis [13, 14]. Besides, MR analyses are not prone to reverse causation as genetic variants are fixed and do not change over time [15]. This makes MR an ideal approach for inferring causality of an exposure on an outcome. However, for MR estimates to be valid, three main assumptions must be met (i) instrumental variables must be strongly associated with the exposure (ii) instrument variables are not associated with other confounders of the exposure and outcome association and (iii) there is no horizontal pleiotropy between the instrument variables and outcome variable [16].

Here we performed a two-sample MR to examine the causal associations between chronic hepatitis infection and risk of extrahepatic cancers using genetic variants associated with chronic HBV infection and chronic HCV infection as instrumental variables. Genetic variants associated with chronic HBV infection and chronic HCV infection were identified from a recent genome-wide association study (GWAS) [17]. We examined the causal associations of genetic variants with risks of the biliary tract, cervical, colorectal, endometrial, esophageal, gastric, liver, lung, ovarian and pancreatic cancers.

## Methods

### Exposure data

Instrumental variables for chronic hepatitis infection were selected from a recent large GWAS with more than 200, 000 individuals of East Asian descent in Japan [17]. A total of 1,394 chronic HBV patients and 5,794 chronic HCV patients were used in the analysis. Independent single nucleotide polymorphisms (SNP) were defined when it was one megabase way from the lead SNP. A total of six and four conditionally independent SNPs were identified to be strongly associated with chronic HBV and chronic HCV infection, respectively at the genome-wide significance threshold (P-value < 5 × 10^−8^). Detailed information for the genetic variants used in this analysis is provided in Table.S1.

### Outcome data

Summary level data for the associations of genetic variants and risks of biliary tract cervical, colorectal, endometrial, esophageal, gastric, liver, lung, ovarian and pancreatic cancers were obtained from GWAS of the East-Asians ancestry [17]. All participants included in the analysis were of East-Asian descent from Japan recruited into the Biobank Japan (BBJ) [18], which was launched in 2003 to provide evidence for the implementation of personalized medicine in Japan by investigating the role of environmental and genetic factors associated with various diseases and phenotypes [18]. The BBJ has over 47 diseases and phenotypes including 13 site-specific cancers.

### Statistical analysis

Our analyses were performed using the simple median method, weighted median method, and MR-Egger method and inverse-variance weighted (IVW) method. The IVW was our primary MR method and it provides accurate estimates when there is no heterogeneity and directional pleiotropy between the exposure and outcome [19]. We performed a two-sample MR using the fixed-effect IVW method. The heterogeneity of causal association between chronic hepatitis infection and extrahepatic cancers were investigated by estimating Cochran’s Q statistics and I^2^ statistics assuming a fixed-effect model [20].

We performed sensitivity analyses to check and correct for the presence of pleiotropy in the causal estimates [19]. We defined the presence of heterogeneity if the Q statistics were significant at a P-value ≤ 0.05 and consequently, we used a random-effect IVW method in our analysis [21]. We assessed the presence of horizontal pleiotropy using MR-Egger regression based on its intercept terms and Mendelian randomisation pleiotropy residual sum and outlier (MR-PRESSO) [22, 23]. When the MR-Egger intercept deviates from zero or its P-value is statistically significant at P-value ≤ 0.05 it indicates the presence of horizontal pleiotropy in the MR analysis [24] and a different MR method was used to report the results. In this analysis, we used the weighted median method in the presence of heterogeneity and horizontal pleiotropy. The weighted median methods can give valid estimates under the presence of horizontal pleiotropy when up to 50% of the instruments variables are valid [19]. MR-PRESSO was used to detect and remove outlier instrumental variable in the MR analysis [23]. Additionally, we performed a multivariable MR analysis using the fixed-effect IVW method with chronic HBV infection adjusted for chronic HCV infection and chronic HCV infection adjusted for chronic HBV infection [25]. All the statistical analyses were performed using the Mendelian Randomisation [26] and Two Sample MR [27] packages in the R programming language.

### Multiple testing correction

We performed multiple testing using the Bonferroni correction method [28]. Since we used six instrumental variables in the MR analysis for chronic HBV infection and extrahepatic cancers and to adjust for multiple testing, our statistically significant P-value was ≤0.0083. For chronic HCV infection, we used four instrumental variables and an adjusted P-value ≤ 0.0125 was used for determining statistically significant results of the causal association between chronic HCV infection and extrahepatic cancers.

## Results

We identified six and four conditionally independent SNPs that were strongly associated with chronic HBV infection and chronic HCV infection, respectively from the BBJ (Table.S1). We defined the SNP to be independent if it were one megabase away from the lead SNP. Notably, most of these SNPs are mapped on chromosome 6 except rs8113007, which is mapped on chromosome 19.

### Chronic HBV infection MR estimates

The IVW was our primary MR method in the absence of horizontal pleiotropy with MR-Egger regression intercept P-value > 0.05 (Table.S2). Firstly, we evaluated whether chronic HBV infection is causally associated with extrahepatic cancers. We found chronic HBV infection to be causally associated with a 34% higher risk of liver cancer, odds ratio (OR) =1.34 and 95% confidence interval (CI) = 1.17-1.53, P-value = 0.007 (Table 1, Fig.1G). Additionally, chronic HBV infection was causally associated with extrahepatic cancers including gastric cancer (OR = 1.19, 95% CI = 1.13-1.25, P-value = 0.001, Fig.1F) and lung cancer (OR = 1.21, 95%CI = 1.14-1.28, P-value = 0.001, Fig.1H). Moreover, we found chronic HBV infection to be positively associated with esophageal cancer (OR = 1.12, 95% CI = 1.01-1.25, P-value = 0.039), however after correction for multiple testing our results were not statistically significant at P-value ≤ 0.0083 (Table 1).

**Table 1.** Mendelian randomization estimates for the causal association between chronic hepatitis infections with site-specific cancers.

| Cancer sites | IVW method |  | Simple median method |  | Weighted median method |  | MR-Egger method |  |
| --- | --- | --- | --- | --- | --- | --- | --- | --- |
|  | OR (95%CI) | P-value | OR (95%CI) | P-value | OR (95%CI) | P-value | OR (95%CI) | P-value |
| **Hepatitis B** |  |  |  |  |  |  |  |  |
| Biliary tract | 1.05 (0.86-1.28) | 0.633 | 1.03 (0.79-1.34) | 0.813 | 1.01 (0.80-1.28) | 0.944 | 1.08 (0.51-2.29) | 0.835 |
| Cervix | 1.20 (0.93-1.55) | 0.153 | **1.41 (1.14-1.76)** | **0.002** | 1.23 (1.02-1.48) | 0.032 | 0.70 (0.27-1.79) | 0.455 |
| Colorectum | 1.04 (0.98-1.10) | 0.209 | 1.04 (0.98-1.10) | 0.212 | 1.04 (0.98-1.10) | 0.170 | 1.02 (0.80-1.30) | 0.865 |
| Endometrium | 1.00 (0.87-1.15) | 0.969 | 1.03 (0.88-1.20) | 0.741 | 1.04 (0.90-1.19) | 0.632 | 0.70 (0.44-1.11) | 0.129 |
| Esophagus | 1.12 (1.01-1.25) | 0.039 | 1.17 (1.02-1.35) | 0.027 | 1.09 (0.97-1.24) | 0.162 | 0.82 (0.56-1.19) | 0.297 |
| Gastric | **1.19 (1.13-1.25)** | **0.001** | **1.23 (1.14-1.33)** | **0.001** | **1.21 (1.13-1.29)** | **0.001** | 1.12 (0.91-1.38) | 0.279 |
| Liver | **1.34 (1.17-1.53)** | **0.007** | **1.33 (1.17-1.52)** | **0.007** | **1.40 (1.25-1.56)** | **0.002** | 1.46 (0.83-2.57) | 0.190 |
| Lung | **1.21 (1.14-1.28)** | **0.001** | **1.21 (1.12-1.32)** | **0.001** | **1.20 (1.12-1.29)** | **0.001** | 0.99 (0.80-1.24) | 0.948 |
| Ovary | 0.98 (0.86-1.13) | 0.807 | 0.99 (0.83-1.18) | 0.887 | 0.98(0.84-1.15) | 0.836 | 0.94(0.56-1.57) | 0.803 |
| Pancreas | 1.01 (0.86-1.20) | 0.868 | 1.06 (0.85-1.33) | 0.597 | 1.03(0.84-1.27) | 0.786 | 0.67(0.35-1.28) | 0.226 |
| **Hepatitis C** |  |  |  |  |  |  |  |  |
| Biliary tract | 0.98 (0.59-1.61) | 0.924 | 0.98 (0.53-1.82) | 0.959 | 0.89 (0.50-1.58) | 0.685 | 0.70 (0.22-2.29) | 0.561 |
| Cervix | 1.24 (0.74-2.06) | 0.411 | 1.58 (0.97-2.58) | 0.069 | 1.15 (0.72-1.84) | 0.555 | 0.80 (0.22-2.97) | 0.742 |
| Colorectum | 1.08 (0.90-1.29) | 0.434 | 1.18 (0.99-1.41) | 0.066 | 1.09 (0.94-1.27) | 0.266 | 0.79 (0.61-1.03) | 0.081 |
| Endometrium | 0.92 (0.69-1.23) | 0.563 | 0.90 (0.63-1.30) | 0.585 | 0.96 (0.68-1.34) | 0.797 | 1.15 (0.58-2.29) | 0.683 |
| Esophagus | 1.11 (0.86-1.43) | 0.425 | 1.22 (0.88-1.68) | 0.230 | 1.12 (0.83-1.52) | 0.472 | 1.03 (0.56-1.89) | 0.926 |
| Gastric | 1.11 (0.98-1.24) | 0.093 | 1.10 (0.95-1.27) | 0.182 | 1.09 (0.95-1.26) | 0.207 | 1.05 (0.77-1.44) | 0.765 |
| Liver | **2.75 (2.21-3.42)** | **0.0008** | **2.83 (2.06-3.88)** | **0.0007** | **2.61 (1.96-3.49)** | **0.0005** | **2.24 (1.33-3.75)** | **0.002** |
| Lung | 1.14 (0.94-1.39) | 0.188 | 1.12 (0.90-1.39) | 0.297 | 1.06 (0.88-1.27) | 0.566 | 0.91 (0.58-1.42) | 0.674 |
| Ovary | 1.19 (0.66-2.15) | 0.566 | 1.39 (0.81-2.39) | 0.238 | 1.18 (0.75-1.87) | 0.474 | 0.47 (0.18-1.19) | 0.110 |
| Pancreas | 1.09 (0.70-1.68) | 0.709 | 1.08 (0.64-1.84) | 0.771 | 1.06 (0.64-1.75) | 0.823 | 0.90 (0.32-2.53) | 0.843 |

IVW; inverse –variance weighted methods, MR; Mendelian randomization, OR; odds ratio, CI; confidence interval

**Fig.1.**
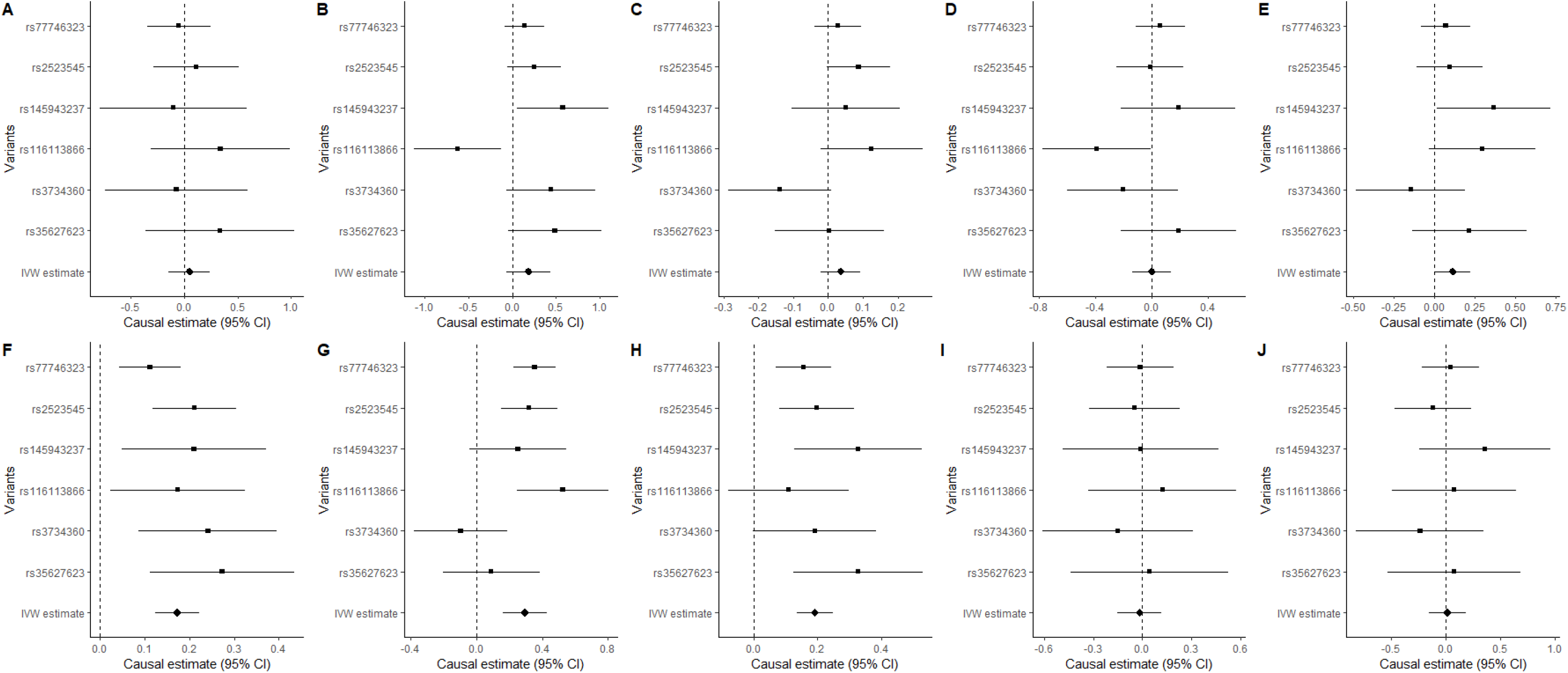
Mendelian randomization of chronic HBV infection and site-specific cancers. (A) biliary tract (B) cervix, (C) colorectum, (D) endometrium, (E) oesophagus, (F) gastric, (G) liver, (H) lung, (I) ovary, and (J) pancreas.

### Chronic HCV infection MR estimates

For this analysis, we only found chronic HCV infection to be causally associated with liver cancer (OR = 2.75, 95%CI = 2.21-3.42, P-value = 0.0008, Fig.2G). Besides IVW method, simple median method (OR = 2.83, 95%CI = 2.06-3.88, P-value = 0.0009), weighted median method (OR = 2.61, 95%CI = 1.96-3.49, P-value = 0.001) and MR-Egger method (OR = 2.24, 95%CI = 1.33-3.75, P-value = 0.002) all show consistent causal association between chronic HCV infection and liver cancer (Table 1).

**Fig.2.**
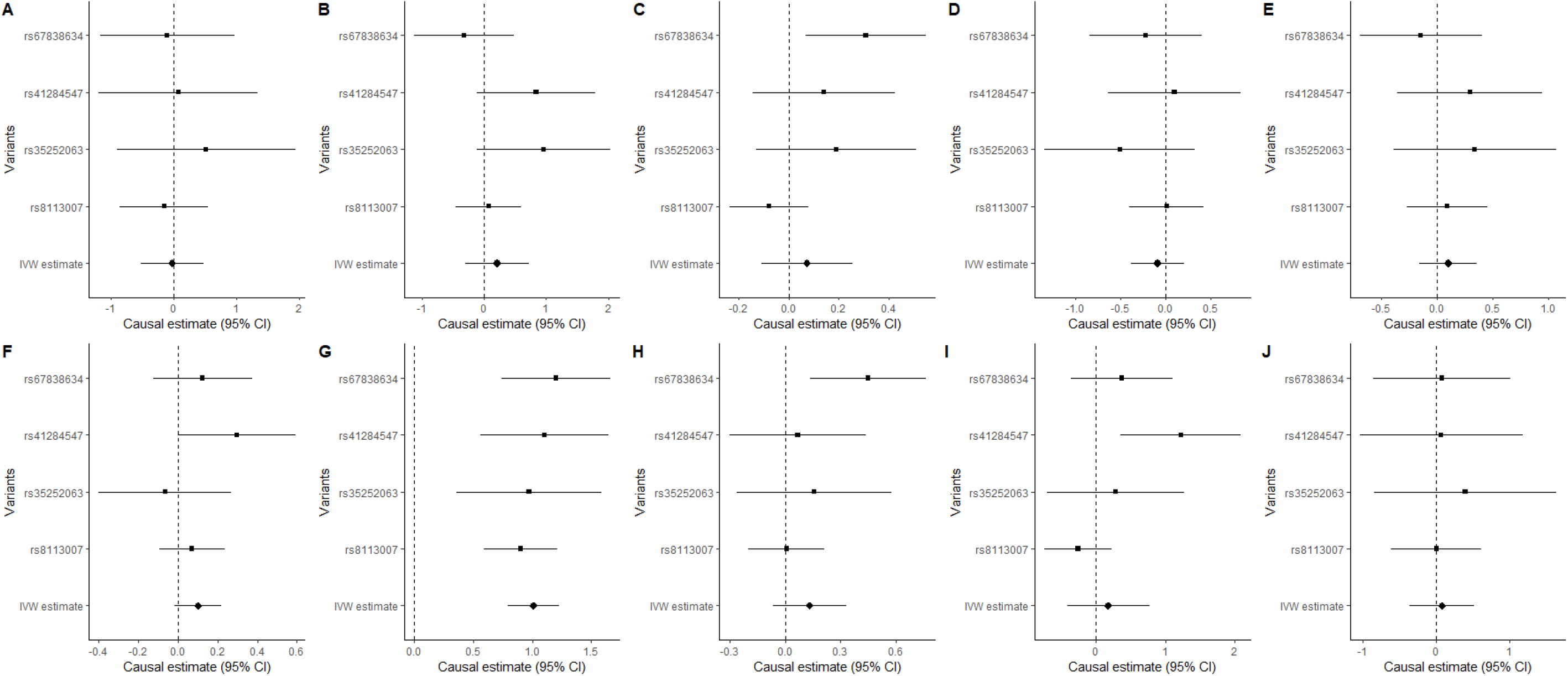
Mendelian randomization of chronic HCV infection and site-specific cancers. (A) biliary tract (B) cervix, (C) colorectum, (D) endometrium, (E) oesophagus, (F) gastric, (G) liver, (H) lung, (I) ovary, and (J) pancreas.

### Sensitivity analyses

We performed sensitivity tests using MR-PRESSO and multivariable MR analysis. We found no evidence of horizontal pleiotropy for chronic HBV infection with all site-specific cancers with P-values >0.05 for the MR-Egger regression intercept approach (Table.S2). However, we found evidence of horizontal pleiotropy for chronic HCV infection with colorectal cancer and ovarian cancer with MR-Egger regression intercepts P-value = 0.012 and 0.030, respectively, (Table.S2). Moreover, we found the presence of heterogeneity for the causal association of chronic HBV infection with cervical cancer (I^2^ = 66.2%, P_het_ =0.011) and also with liver cancer (I^2^ = 60.9%, P_het_ =0.025, Table.S2). For chronic HCV infection, we found evidence for high heterogeneity for colorectal cancer (I^2^ =62.4%, P_het_ = 0.046) and also in ovarian cancer (I^2^ = 66.5%, P_het_ =0.029, Table.S2).

We then performed a sensitivity analysis and our results revealed rs116113866 as a genetic instrument driving the null association between chronic HBV infection and cervical cancer (OR = 1.04, 95%CI = 0.98-1.10, P-value = 0.209, Fig.1B). However, when we performed MR-PRESSO, which identify and remove outlier instrumental variables in MR analysis. Our outlier corrected MR-PRESSO for chronic HBV infection was causally associated with cervical cancer (OR =1.30, 95%CI = 1.12-1.51, P-value = 0.0263). However, this finding was not statistically significant after Bonferroni corrections. Interestingly, for chronic HBV infection and liver cancer, our MR-PRESSO analysis was similar to the IVW method regardless of the presence of heterogeneity, which was corrected by using the random IVW method. Since there were heterogeneity and horizontal pleiotropy for chronic HCV infection with colorectal cancer, and ovarian cancer (Table.S2), we, therefore, used the weighted median method for MR analysis, which gives valid estimates under the presence of horizontal pleiotropy when up to 50% of the instruments variables are valid. However, our weighted median MR analysis for colorectal cancer (OR =1.09, 95%CI = 0.94-1.27. P-value = 0.266) and for ovarian cancer (OR =1.18, 95%CI = 0.75-1.87, P-value =0.474) were not statistically significant (Table 1).

We further performed the multivariable MR analyses adjusting for chronic HCV infection and our results were largely unchanged from our main IVW results. Similarly, adjusting for chronic HBV infection did not attenuate the statistically significant results of chronic HCV infection and liver cancer.

## Discussion

Our MR study was set out to investigate whether chronic hepatitis infection was causally associated with extrahepatic cancers. We found chronic HBV infection to be causally associated with extrahepatic cancers including gastric and lung cancer. Additionally, chronic HBV infection and chronic HCV infection were all causally associated with liver cancer, supporting the overwhelming evidence from observational studies that chronic hepatitis infection is a risk factor for liver cancer [3–6].

Using the IVW method, we found chronic HBV infection to be causally associated with gastric cancer, corroborating previous observational studies [7, 29]. However, our previous analysis using the Taiwan National Health Insurance datasets failed to find any significant association between chronic HBV infection and gastric cancer [11]. In this MR analysis, only chronic HBV infection was significantly associated with gastric cancer suggesting that HBV may play a crucial role in tumorigenesis of gastric cancer. Although we found chronic HCV infection not to be significantly associated with gastric cancer (OR =1.11, 95% CI = 0.98-1.24, P-value = 0.093), the odds of developing gastric cancer was high among patients with chronic HCV infection (Table 1). Observational studies reported a high risk of developing lung cancer among patients with chronic HBV infection [6, 30], supporting our MR analysis that chronic HBV infection is a risk factor for lung cancer. HBV a partially circular double-stranded DNA virus encode HBV protein X, a transcriptional coactivator that plays a pivotal role in tumorigenesis by modulating key regulators of the apoptosis, interfering with the DNA repair pathways and tumour suppressor genes [31].

Prior evidence indicated that chronic HBV infection is associated with biliary tract, esophageal, endometrial, pancreatic, and colorectal cancer [6, 7, 9, 11, 30]. Although, we failed to find any significant association between chronic HBV infection and these site-specific cancers, our ORs for these associations were >1 (Table 1), suggesting that chronic HBV infection may be a risk factor for these site-specific cancers. Surprisingly, we found chronic HBV infection to be inversely associated with ovarian cancer, which is contrary to our previous analysis in Han Chinese patients [11]. However, this association was not statistically significant and further validations are needed by future studies.

Chronic HCV infection was causally associated with liver cancer, supporting the overwhelming evidence of a well-known causal association between chronic HCV infection and liver cancer from previous observational studies [3–5]. However, our MR analysis failed to found any significant association between chronic HCV infection and extrahepatic cancers, which is contrary to our previous analysis indicating that chronic HCV infection is a risk factor for ovarian, non-Hodgkin’s lymphoma, gallbladder and extrahepatic bile duct cancer [11]. In this analysis, we did not have summary level data for non-Hodgkin’s lymphoma, gallbladder and extrahepatic bile duct cancer, limiting our ability to assess this association further. Nevertheless, chronic HCV infection was not significantly associated with ovarian cancer, but our OR was 1.19 thus suggesting a possible positive association between chronic HCV infection and ovarian cancer which requires further follow up. Moreover, chronic HCV infection was not significantly associated with the biliary tract and endometrial cancer, suggesting that chronic HCV infection may not be a risk factor for these site-specific cancers.

Our study has several strengths include the use of the MR approach, which eliminate some confounders commonly observed in epidemiological studies. Moreover, we used multiple SNPs, which were strongly associated with chronic hepatitis HBV and chronic HCV infection. Besides, we use a homogenous population which minimises heterogeneity commonly observed when individuals of different ancestry population are used in genetic studies. However, our study had some limitations including a lack of power to find a significant association between chronic hepatitis infection and certain site-specific cancers. Our analyses included individuals of East Asian descent where there is a high prevalence of chronic hepatitis infection and therefore our results might not be generalizable to other ancestry populations. Another limitation is the possibility of having overlapping individuals in predictor and outcome summary statistics, which might introduce some bias in our MR estimation. Nevertheless, we performed a sensitivity analysis using four different approaches and our results were consistent and robust.

## Conclusions

In summary, our MR results support a well-established relationship between chronic hepatitis infection and liver cancer and suggest that chronic HBV infection may also be a risk factor for gastric cancer and lung cancer. However, we found no evidence for the causal association between chronic HCV infection and extrahepatic cancers and future studies should investigate this further.

## Data Availability

GWAS summary statistics supporting the findings of this study are available at Biobank Japan

https://humandbs.biosciencedbc.jp/en/hum0014-v19#41diseases

## Declarations

### Authors’ contributions

Conceptualization, ABK; methodology, ABK, FS, CCY, TC writing-original draft preparation, ABK, FS, TC; writing-review and editing, ABK, FS, MGS, CCY, TC.

### Consent for publication

Not applicable

### Availability of data and materials

GWAS summary statistics supporting the findings of this study are available at https://humandbs.biosciencedbc.jp/en/hum0014-v19#41diseases

### Ethics approval and consent to participate

The Biobank Japan obtained informed consent from the study participants and approval from its institutional review board.

## Acknowledgements

The study has been supported by the South African Medical Research Council (with funds received from the South African National Department of Health) and the UK Medical Research Council (with funds from the UK Government’s Newton Fund) (MRC-RFA-SHIP 01-2015) for the Evolving Risk Factors for Cancers in African populations study (ERICA-SA). The funders stated therein had no role in the paper design, data collection, data analysis, interpretation and writing of the paper. TC is an international training fellow supported by the Wellcome Trust grant (214205/Z/18/Z). The contents of this publication are solely the responsibility of the authors and do not necessarily represent the official views of the South African Medical Research Council or the South African National Department of Health or the UK Medical Research Council from the UK Government’s Newton Fund.

## Supplementary material

**Table S1.** Instrumental variables used in the causal analysis of chronic hepatitis infection with various site-specific cancers.

| SNP | Chr | Position | A1 | A2 | MAF | Beta | SE | Nearest genes | P-value |
| --- | --- | --- | --- | --- | --- | --- | --- | --- | --- |
| **Hepatitis B** |  |  |  |  |  |  |  |  |  |
| rs77746323 | 6 | 33036401 | C | T | 0.599 | 0.517 | 0.040 | *HLA-DPA1* | $1.32 \times 10^{-38}$ |
| rs2523545 | 6 | 31333499 | G | A | 0.872 | 0.551 | 0.060 | *DHFRP2* | $5.61 \times 10^{-20}$ |
| rs145943237 | 6 | 26865972 | G | T | 0.725 | 0.271 | 0.048 | *GUSBP2* | $1.18 \times 10^{-08}$ |
| rs116113866 | 6 | 30286105 | C | G | 0.043 | -0.582 | 0.102 | *HCG17* | $1.34 \times 10^{-08}$ |
| rs3734360 | 6 | 34100568 | G | A | 0.033 | -0.640 | 0.116 | *GRM4* | $3.36 \times 10^{-08}$ |
| rs35627623 | 6 | 28583377 | A | C | 0.885 | 0.355 | 0.065 | *SCAND3* | $4.54 \times 10^{-08}$ |
| **Hepatitis C** |  |  |  |  |  |  |  |  |  |
| rs67838634 | 6 | 32662128 | A | G | 0.437 | -0.142 | 0.019 | *MTCO3P1* | $8.11 \times 10^{-14}$ |
| rs41284547 | 6 | 31390139 | T | A | 0.557 | 0.128 | 0.020 | *HCP5* | $3.11 \times 10^{-10}$ |
| rs35252063 | 6 | 29606954 | A | G | 0.279 | -0.119 | 0.021 | *SUMO2P1* | $2.26 \times 10^{-08}$ |
| rs8113007 | 19 | 39743103 | A | T | 0.105 | 0.356 | 0.033 | *MSRB1P1* | $6.59 \times 10^{-28}$ |

SNP; single nucleotide polymorphisms, Chr: chromosome, MAF; minor allele frequency, SE; standard error

**Table S2.**
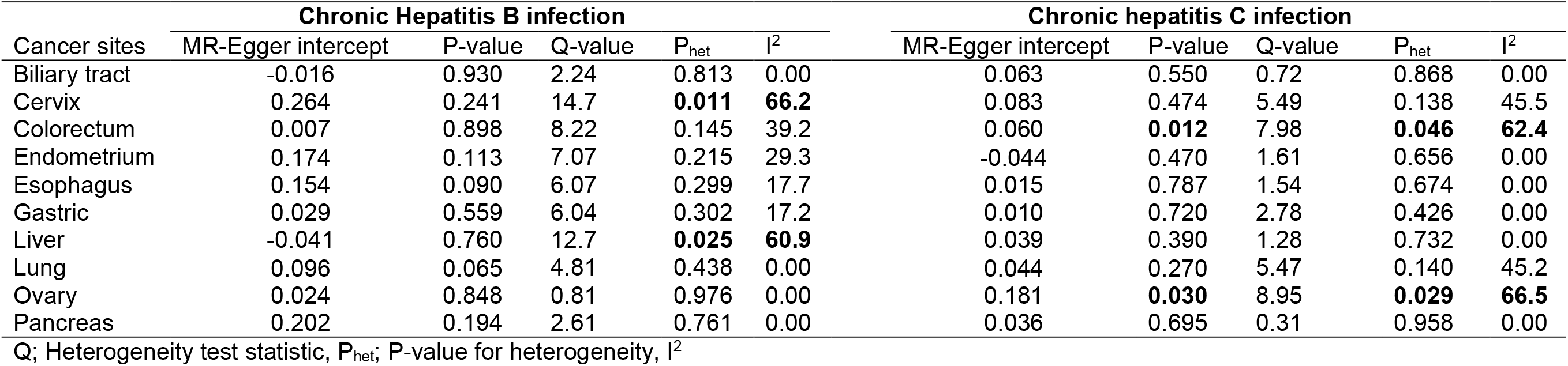
Assessment of pleiotropy for the association between chronic hepatitis infections with site-specific cancers.

## References

1. Razavi-Shearer D, Gamkrelidze I, Nguyen MH, et al (2018) Global prevalence, treatment, and prevention of hepatitis B virus infection in 2016: a modelling study. The Lancet Gastroenterology & Hepatology 3:383–403

2. Hajarizadeh B, Grebely J, Dore GJ (2013) Epidemiology and natural history of HCV infection. Nat Rev Gastroenterol Hepatol 10:553–562

3. Hadziyannis S, Tabor E, Kaklamani E, Tzonou A, Stuver S, Tassopoulos N, Mueller N, Trichopoulos D (1995) A case-control study of hepatitis B and C virus infections in the etiology of hepatocellular carcinoma. Int J Cancer 60:627–631

4. Yu MC, Tong MJ, Coursaget P, Ross RK, Govindarajan S, Henderson BE (1990) Prevalence of hepatitis B and C viral markers in black and white patients with hepatocellular carcinoma in the United States. J Natl Cancer Inst 82:1038–1041

5. Perz JF, Armstrong GL, Farrington LA, Hutin YJF, Bell BP (2006) The contributions of hepatitis B virus and hepatitis C virus infections to cirrhosis and primary liver cancer worldwide. J Hepatol 45:529–538

6. Sundquist K, Sundquist J, Ji J (2014) Risk of hepatocellular carcinoma and cancers at other sites among patients diagnosed with chronic hepatitis B virus infection in Sweden. J Med Virol 86:18–22

7. Song C, Lv J, Liu Y, et al (2019) Associations Between Hepatitis B Virus Infection and Risk of All Cancer Types. JAMA Netw Open. https://doi.org/10.1001/jamanetworkopen.2019.5718

8. Amin J, Dore GJ, O’Connell DL, Bartlett M, Tracey E, Kaldor JM, Law MG (2006) Cancer incidence in people with hepatitis B or C infection: a large community-based linkage study. J Hepatol 45:197–203

9. Tian T, Song C, Jiang L, et al (2020) Hepatitis B virus infection and the risk of cancer among the Chinese population. International Journal of Cancer 147:3075–3084

10. Hong CY, Sinn DH, Kang D, Paik SW, Guallar E, Cho J, Gwak G-Y (2020) Incidence of extrahepatic cancers among individuals with chronic hepatitis B or C virus infection: A nationwide cohort study. Journal of Viral Hepatitis 27:896–903

11. Kamiza AB, Su F-H, Wang W-C, Sung F-C, Chang S-N, Yeh C-C (2016) Chronic hepatitis infection is associated with extrahepatic cancer development: a nationwide population-based study in Taiwan. BMC Cancer 16:

12. Davey Smith G, Hemani G (2014) Mendelian randomization: genetic anchors for causal inference in epidemiological studies. Hum Mol Genet 23:R89–98

13. Davey Smith G, Ebrahim S (2003) ‘Mendelian randomization’: can genetic epidemiology contribute to understanding environmental determinants of disease?*. International Journal of Epidemiology 32:1–22

14. Lawlor DA, Harbord RM, Sterne JAC, Timpson N, Davey Smith G (2008) Mendelian randomization: using genes as instruments for making causal inferences in epidemiology. Stat Med 27:1133–1163

15. Sekula P, Del Greco M F, Pattaro C, Köttgen A (2016) Mendelian Randomization as an Approach to Assess Causality Using Observational Data. J Am Soc Nephrol 27:3253– 3265

16. Bowden J, Davey Smith G, Haycock PC, Burgess S (2016) Consistent Estimation in Mendelian Randomization with Some Invalid Instruments Using a Weighted Median Estimator. Genet Epidemiol 40:304–314

17. Ishigaki K, Akiyama M, Kanai M, et al (2020) Large-scale genome-wide association study in a Japanese population identifies novel susceptibility loci across different diseases. Nature Genetics 52:669–679

18. Nagai A, Hirata M, Kamatani Y, et al (2017) Overview of the BioBank Japan Project: Study design and profile. J Epidemiol 27:S2–S8

19. Burgess S, Bowden J, Fall T, Ingelsson E, Thompson SG (2017) Sensitivity Analyses for Robust Causal Inference from Mendelian Randomization Analyses with Multiple Genetic Variants. Epidemiology 28:30–42

20. Higgins JPT, Thompson SG, Deeks JJ, Altman DG (2003) Measuring inconsistency in meta-analyses. BMJ 327:557–560

21. Bowden J, Del Greco M F, Minelli C, Davey Smith G, Sheehan N, Thompson J (2017) A framework for the investigation of pleiotropy in two-sample summary data Mendelian randomization. Stat Med 36:1783–1802

22. Bowden J, Davey Smith G, Burgess S (2015) Mendelian randomization with invalid instruments: effect estimation and bias detection through Egger regression. Int J Epidemiol 44:512–525

23. Verbanck M, Chen C-Y, Neale B, Do R (2018) Detection of widespread horizontal pleiotropy in causal relationships inferred from Mendelian randomization between complex traits and diseases. Nat Genet 50:693–698

24. Hemani G, Bowden J, Davey Smith G (2018) Evaluating the potential role of pleiotropy in Mendelian randomization studies. Human Molecular Genetics 27:R195–R208

25. Burgess S, Thompson SG (2015) Multivariable Mendelian randomization: the use of pleiotropic genetic variants to estimate causal effects. Am J Epidemiol 181:251–260

26. Yavorska OO, Burgess S (2017) MendelianRandomization: an R package for performing Mendelian randomization analyses using summarized data. Int J Epidemiol 46:1734–1739

27. Hemani G, Zheng J, Elsworth B, et al (2018) The MR-Base platform supports systematic causal inference across the human phenome. Elife. https://doi.org/10.7554/eLife.34408

28. Benjamini Y, Hochberg Y (1995) Controlling the False Discovery Rate: A Practical and Powerful Approach to Multiple Testing. Journal of the Royal Statistical Society Series B (Methodological) 57:289–300

29. Wei X-L, Qiu M-Z, Jin Y, et al (2015) Hepatitis B virus infection is associated with gastric cancer in China: an endemic area of both diseases. Br J Cancer 112:1283– 1290

30. An J, Kim JW, Shim JH, et al (2018) Chronic hepatitis B infection and non-hepatocellular cancers: A hospital registry-based, case-control study. PLOS ONE 13:e0193232

31. Gómez-Moreno A, Garaigorta U (2017) Hepatitis B Virus and DNA Damage Response: Interactions and Consequences for the Infection. Viruses. https://doi.org/10.3390/v9100304

